# Occupational Farm Work Activities Influence Workers’ Indoor Home Microbiome

**DOI:** 10.1101/2023.08.17.23293194

**Authors:** Kathryn R. Dalton, Mikyeong Lee, Ziyue Wang, Shanshan Zhao, Christine G. Parks, Laura E. Beane-Freeman, Alison A. Motsinger-Reif, Stephanie J. London

## Abstract

**Background:** Farm work entails a heterogeneous mixture of exposures that vary considerably across farms and farmers. Farm work is associated with various health outcomes, both adverse and beneficial. One mechanism by which farming exposures can impact health is through the microbiome, including the indoor built environment microbiome. It is unknown how individual occupational exposures shape the microbial composition in workers’ homes.

**Objectives:** We investigated associations between farm work activities, including specific tasks and pesticide use, and the indoor microbiome in the homes of 468 male farmers.

**Methods:** Participants were licensed pesticide applicators, mostly farmers, enrolled in the Agricultural Lung Health Study from 2008-2011. Vacuumed dust from participants’ bedrooms underwent whole-genome shotgun sequencing for indoor microbiome assessment. Using questionnaire data, we evaluated 6 farm work tasks (processing of either hay, silage, animal feed, fertilizer, or soy/grains, and cleaning grain bins) and 19 pesticide ingredients currently used in the past year, plus 7 persistent banned pesticide ingredients ever used.

**Results:** All 6 work tasks were associated with increased within-sample microbial diversity, with a positive dose-response for the sum of tasks (p=0.001). All tasks were associated with altered overall microbial compositions (weighted UniFrac p=0.001) and with higher abundance of specific microbes, including soil-based microbes such as *Haloterrigena*. Among the 19 pesticides, only current use of glyphosate and past use of lindane were associated with increased within-sample diversity (p=0.02-0.04). Ten currently used pesticides and all 7 banned pesticides were associated with altered microbial composition (p=0.001-0.04). Six pesticides were associated with differential abundance of certain microbes.

**Discussion:** Specific farm activities and exposures can impact the dust microbiome inside homes. Our work suggests that occupational farm exposures could impact the health of workers and their families through modifying the indoor environment, specifically the microbial composition of house dust, offering possible future intervention targets.

## Introduction

Farm work is a demanding occupation. Farmers have long been recognized as being at high risk of injury, respiratory diseases (e.g., farmers’ lung), dermal conditions (e.g., irritant dermatitis), and certain cancers.(1–4) Farm work is not consistent across workers as it requires individuals to carry out a variety of tasks. Farmers, hired farmworkers, and farm family members may operate agricultural machinery, handle crops and livestock, build and repair equipment, and apply agrochemicals such as pesticides and fertilizers, which may put them at risk of some diseases. If farm type were the sole explanation for the morbidity pattern among farmers, we might expect disease prevalence and incidence to be quite similar among all farmers engaging in the same type of farming, but this does not appear to be the case.(5–7) This suggests that farming entails a heterogeneous mixture of individualized tasks and exposures, and points to the need for careful evaluation of the farm environment.

Multiple theories have been offered to understand how farming exposures exert biological effects that lead to respiratory and other disease conditions. One of the most supported by the literature is through the microbiome, the collection of microorganisms within a single site, and its link to immune function.(8) Several studies have shown altered host microbiome – gut, nasal, oral, and skin – related to farm work.(9–11) Yet, it is not only the microbes in our bodies that cause biological effects. Humans contribute and are exposed to environmental microbes, especially indoors where they spend the most time.(12) The home built environment microbiome can influence health outcomes by altering the human host microbiome, as well as through direct effects on biologic processes (13), and is also associated with allergic, atopic, and respiratory conditions.(14–17)

The indoor microbiota can be influenced by environmental factors, such as farming. The authors and others have previously shown altered home dust microbiota with living on a farm and by farm type.(18–20) However, it is unknown how different occupational tasks and exposures within farm work shape the microbes in workers’ homes, which is important for their health and the health of cohabitants. There is some evidence that the host microbiome is impacted by pesticides and other chemicals used by farmers and other occupational groups.(21–23) Yet, none of these studies have looked at the effect on built environment microbes. Therefore, we evaluated whether farm work activities and pesticide use were associated with differences in the indoor dust microbiome in the homes of 468 male participants of the Agricultural Lung Health Study.

## Methods

### Study Design & Population

The Agricultural Lung Health Study (ALHS) is a case-control study of asthma nested within the Agricultural Health Study (AHS), a prospective cohort of licensed private pesticide applicators, mostly farmers, and their spouses from Iowa (IA) and North Carolina (NC), USA.(24) (ALHS data release P3REL201209.00) Further details on the study design and inclusion criteria can be found elsewhere.(18, 19) Of the full ALHS cohort (N=3301), 2871 participants completed a home visit between 2009 to 2013 with vacuumed bedroom dust collection.(25) The Institutional Review Board at the National Institute of Environmental Health Sciences approved the study. Written informed consent was obtained from all participants.

### Home Dust Microbiome Examination

A trained field technician vacuumed the sleeping surface and a two square yard area (1.68-m^2^) on the floor next to the bed for 4 minutes with a DUSTREAM Collector (Indoor Biotechnologies Inc.). A subset of 879 dust samples were sent for microbiome analysis, with sample selection described elsewhere.(18, 19, 25) DNA extraction followed standard protocols following manufactured kits and is described elsewhere.(18, 19) Extracted DNA samples were sent to the University of California San Diego IGM Genomics Center for library preparation, multiplexing, and whole genome shotgun sequencing using standard techniques.(26) Details on the full library preparation, sequencing protocols, and quality control steps are described in Wang et. al.(19) After quality control, 781 samples remained with 6,528 taxa for downstream analysis. A taxonomy chart was created that assigned all taxa to a taxonomic classification across the seven phylogenetic levels - kingdom, phylum, class, order, family, genus, and species. We filtered out samples with a minimum library size of less than 1,003 base pairs.

### Exposure Assessment

A technician recorded home cleanliness on a standardized five-point scale (27), which was aggregated to a binary variable comprising poor/lower (score of 1-2) or good/higher (score of 3– 5) home condition. We categorized season of dust collection based on the date of the home visit: March 21–June 20 for spring, June 21–September 20 for summer, September 21–December 20 for fall, and December 21–March 20 for winter.

Information on smoking status and indoor furry pets (dogs/cats) was reported on questionnaires. Participants reported whether they had performed the following 6 work tasks within the past 12 months; handled hay, silage (fermented grass and other plants), or soybeans/grains, ground feed for animals, fertilized fields, and/or cleaned grain bins. Participants also provided the names of pesticide products used within the past 12 months (current use). Reported names were linked to pesticide active ingredient names using the Environmental Protection Agency (EPA) Pesticide Classification Code.(28) We restricted analysis to the 19 pesticide active ingredients currently used by at least 10 participants – 13 herbicides and 6 insecticides, which includes aggregated composite pesticide classes pyrethroid and organophosphate. In previous AHS surveys, participants provided names of agrochemicals used ever in their lifetimes. In the current work, we additionally analyzed past use of 7 banned organochlorine insecticides due to their long half-lives, bio-accumulation and/or persistence in the environment. For 11 of the 19 currently used active pesticide ingredients and all 7 banned past use pesticide ingredients, we calculated lifetime days of use, estimated from the average days of use per year and the total years the participant reported using the pesticide active ingredient (information was not available to calculate lifetime days of use for 8 currently used pesticides).

### Statistical Analysis

As we focused on direct exposure to farm work tasks and pesticides, we restricted analyses to dust samples from the 468 male pesticide applicators because 98% of pesticide applicators were male and the frequency of work tasks and pesticide use reported among female spouses (N=313) was lower (29). The primary exposure variables were the 6 self-reported work tasks performed within the past year, treated as binary yes/no variables. Additionally, the total number of reported tasks (0–6) for each participant was assessed as both a discrete categorical variable and, for dose-response effect, an ordinal variable. For the 19 currently used pesticide active ingredients, we compared participants who reported use within the past 12 months to participants who reported never using the specific pesticides in their lifetimes (current vs. never). Thus, individuals who reported only past use of the active ingredient (more than 12 months prior) were not included in the analysis for that active ingredient. For the 7 banned pesticides, any past use of the active pesticide ingredient ever in the participants’ lifetimes was compared to never use (ever vs. never). Lifetime days of use was available for 18 pesticide ingredients and was dichotomized at the median among users, generating three categories for analysis – never use, days of use below median, and days of use above median. We assessed correlation between self-reported work tasks and pesticide use via tetrachoric correlation (30) and Spearman’s correlation (31) for categorical work task total.

We performed all statistical analyses and visualization in R v4.1.2 (32), and estimated diversity indices using phyloseq R package.(33) All models included age (continuous), smoking status (never, former, or current), state of residence (IA or NC), asthma status (case or noncase, see House et. al. for case definition (34)), indoor pets (yes or not present), home condition (high or low), and home visit season (winter, spring, summer, or fall), as these were determined to be confounders *a priori*. To evaluate within-sample alpha diversity and its association with work tasks and pesticide use exposures, we used the Shannon alpha diversity index as the outcome in generalized linear models for each exposure. We ran additional adjusted models to evaluate the independent effect of work tasks and pesticide exposures. To explore beta diversity, we calculated unweighted and weighted UniFrac distance metrics. We conducted permutational multivariate analysis of variance (PERMANOVA) models to test the differences in microbial community structure across exposure groups using the *adonis* method in the R vegan package v2.5.7 (35, 36), which reports the R^2^ value to quantify the percentage of variance explained and the p-value for the F-statistic for compositional heterogeneity by exposure groups. We set p<0.05 as the statistical significance threshold for all alpha and beta diversity analyses. To test differentially abundant taxa by exposure groups, we used analysis of composition of microbiomes with bias correction (ANCOM-BC, v1.0.5) models (37) based on a linear regression framework on the log transformed taxa counts. To account for the influence of sequencing depth on taxa counts, we performed normalization by estimating the sampling fraction using the ANCOM-BC built-in algorithm. We tested taxa at the OTU level and summarized the results by genus rank. The coefficients presented are the log fold-difference of the mean normalization abundance difference by ANCOM-BC across exposure levels. Significantly differentially taxa were determined by Benjamini-Hochberg false discovery rate (FDR) controlled p-value of <0.05.

## Results

### Study Population Characteristics and Exposures

**Table 1** summarizes the demographic characteristics and environmental exposures of the study population. Seventy percent of the study population was from Iowa. Participants had a median age of 61 years (IQR 15). Most participants (58%) were never smokers and 33% were asthma cases. Thirty eight percent had indoor pets and 78% had good/higher home condition. Home visits were roughly evenly split across the four seasons (from 17% to 30%).

**Table 1:**
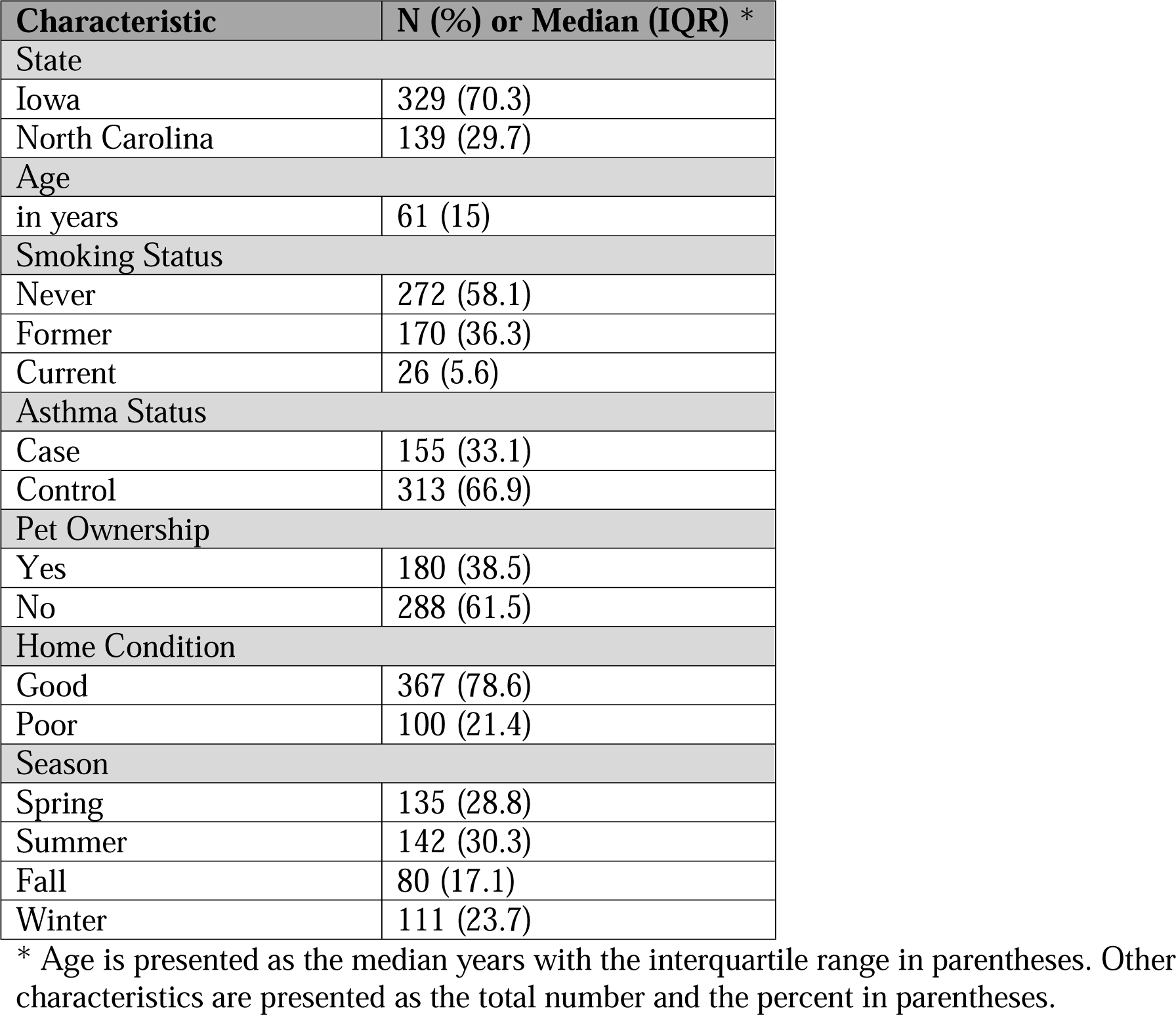
Study Population Characteristics (N=468)

**Table 2** displays the participants’ reported work tasks. The most commonly reported task was handled soy and grains (76%), followed by handled hay (59%), cleaned grain bins (49%), fertilized fields (39%), ground animal feed (27%), and handled silage (17%). The total number of tasks an individual reported, shown in **Table 3**, was evenly spread out from 0 to 6, ranging from 10% (6 tasks) to 18% (0 and 2 tasks).

**Table 2:** Prevalence of Work Tasks in Study Population.

| **Work Tasks** | **N (%) reporting Yes** |
| --- | --- |
| Fertilized Fields | 181 (38.7) |
| Cleaned Grain Bins | 228 (48.7) |
| Handled Hay | 279 (59.6) |
| Handled Silage | 81 (17.3) |
| Ground Animal Feed | 127 (27.1) |
| Handled Soybeans & Grains | 357 (76.3) |

**Table 3:** Distribution of Work Task Totals.

| **Total Work Tasks\*** | **N (%)** |
| --- | --- |
| 0 Tasks | 86 (18.4) |
| 1 Tasks | 63 (13.5) |
| 2 Tasks | 84 (17.9) |
| 3 Tasks | 68 (14.5) |
| 4 Tasks | 63 (13.5) |
| 5 Tasks | 58 (12.4) |
| 6 Tasks | 46 (9.8) |

\* Total work tasks is the individual sum of all work tasks a participant reported Yes to, with a maximum of 6.

**Table 4** shows all 26 pesticide active ingredients with the 19 currently used pesticides listed first. The herbicide glyphosate was the most commonly reported current pesticide (238 users, 86% total), followed by 2,4-D (148, 70%) and atrazine (85, 52%). Among the 7 banned past use pesticides examined, DDT was the most frequently reported (28%), followed by chlordane (27%) and lindane (26%). **Supplemental Table ST1** shows the quantitative lifetime days of use for both current and past use pesticide active ingredients.

**Table 4:**
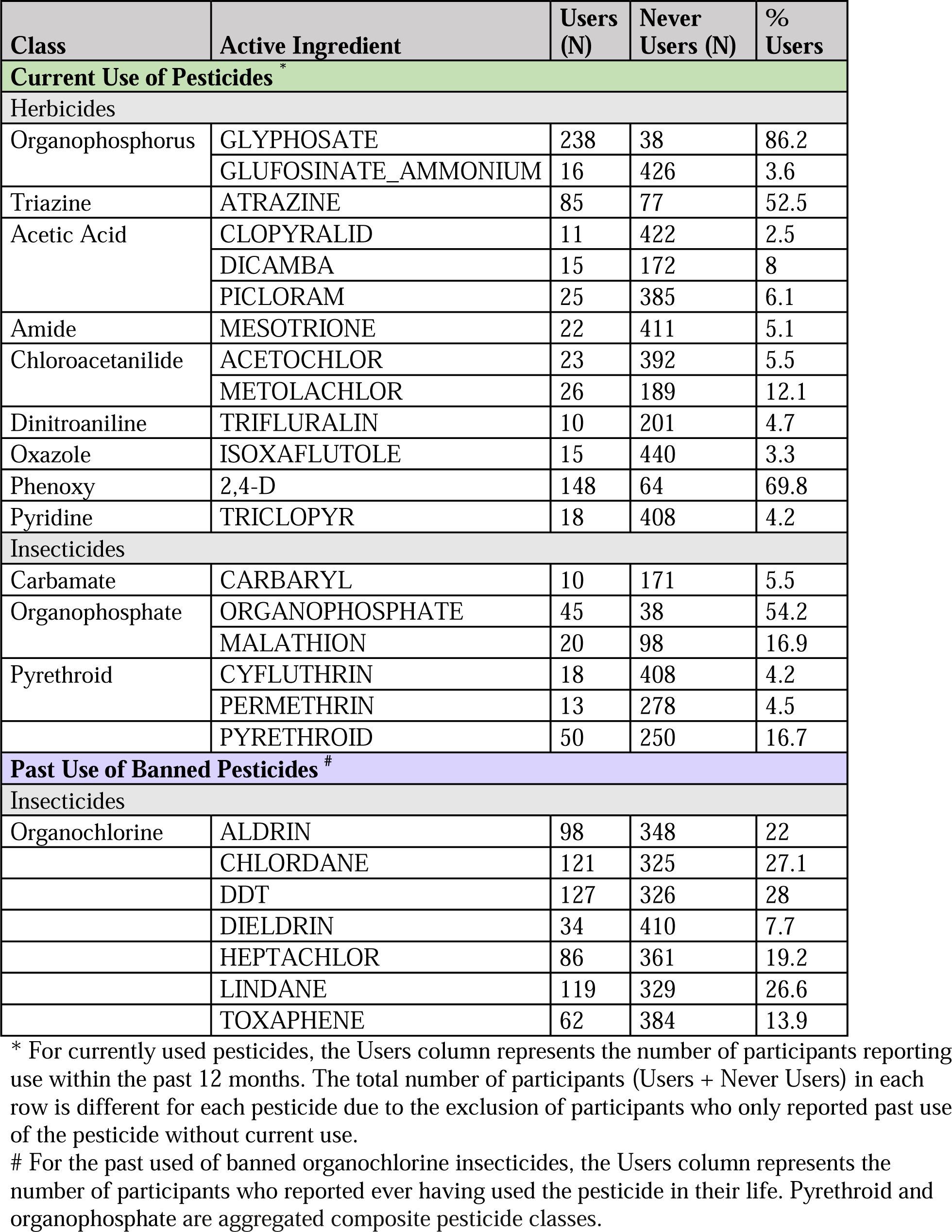
Prevalence of Pesticide Use in Study Population.

| Class | Active Ingredient | Users (N) | Never Users (N) | % Users |
| --- | --- | --- | --- | --- |
| **Current Use of Pesticides \*** |  |  |  |  |
| Herbicides |  |  |  |  |
| Organophosphorus | GLYPHOSATE | 238 | 38 | 86.2 |
|  | GLUFOSINATE_AMMONIUM | 16 | 426 | 3.6 |
| Triazine | ATRAZINE | 85 | 77 | 52.5 |
| Acetic Acid | CLOPYRALID | 11 | 422 | 2.5 |
|  | DICAMBA | 15 | 172 | 8 |
|  | PICLORAM | 25 | 385 | 6.1 |
| Amide | MESOTRIONE | 22 | 411 | 5.1 |
| Chloroacetanilide | ACETOCHLOR | 23 | 392 | 5.5 |
|  | METOLACHLOR | 26 | 189 | 12.1 |
| Dinitroaniline | TRIFLURALIN | 10 | 201 | 4.7 |
| Oxazole | ISOXAFLUTOLE | 15 | 440 | 3.3 |
| Phenoxy | 2,4-D | 148 | 64 | 69.8 |
| Pyridine | TRICLOPYR | 18 | 408 | 4.2 |
| Insecticides |  |  |  |  |
| Carbamate | CARBARYL | 10 | 171 | 5.5 |
| Organophosphate | ORGANOPHOSPHATE | 45 | 38 | 54.2 |
|  | MALATHION | 20 | 98 | 16.9 |
| Pyrethroid | CYFLUTHRIN | 18 | 408 | 4.2 |
|  | PERMETHRIN | 13 | 278 | 4.5 |
|  | PYRETHROID | 50 | 250 | 16.7 |
| **Past Use of Banned Pesticides #** |  |  |  |  |
| Insecticides |  |  |  |  |
| Organochlorine | ALDRIN | 98 | 348 | 22 |
|  | CHLORDANE | 121 | 325 | 27.1 |
|  | DDT | 127 | 326 | 28 |
|  | DIELDRIN | 34 | 410 | 7.7 |
|  | HEPTACHLOR | 86 | 361 | 19.2 |
|  | LINDANE | 119 | 329 | 26.6 |
|  | TOXAPHENE | 62 | 384 | 13.9 |

\* For currently used pesticides, the Users column represents the number of participants reporting use within the past 12 months. The total number of participants (Users + Never Users) in each row is different for each pesticide due to the exclusion of participants who only reported past use of the pesticide without current use.

### Moderate to Strong Correlations Among Works Tasks and Pesticide Use

The correlation between work tasks and pesticide use is presented in **Supplemental Figure SF1**. Correlation to pesticide use was based on current use of 19 active ingredients and ever use of 7 banned active ingredients. Work tasks were highly positively correlated with each other (tetrachoric rho statistic range 0.47–0.87 with only one correlation<0.57), as were past use of banned pesticides (0.30–0.81). Correlations among currently used pesticides ranged from -0.46 to 0.96. A few currently used pesticides (glyphosate, atrazine, 2,4-D, pyrethroid and organophosphate) were moderately correlated with the various work tasks (highest 0.82), with lower correlations for the other pesticides. As expected, banned pesticides had lower correlation with work tasks (-0.39–0.25).

### Work Tasks Strongly Associate with the Home Dust Microbiome

**Figure 1** shows associations between the work tasks and Shannon alpha diversity index. All 6 work tasks were positively associated with within-sample alpha diversity levels (coefficients range from 0.15-0.23, all low 95% confidence intervals >0.045). Due to the strong correlations among the work tasks, it was not possible to determine the independent effect of each one adjusted for all other tasks. Therefore, we focused on evaluating their combined effects with the summed work task variable. A strong positive dose-response pattern was observed for the associations of total number of work tasks and alpha diversity (range of coefficients 0.17 [2 tasks] to 0.45 [6 tasks] compared to no tasks, all low 95% confidence intervals >0.03, p-value for trend <0.001).

**Figure 1:**
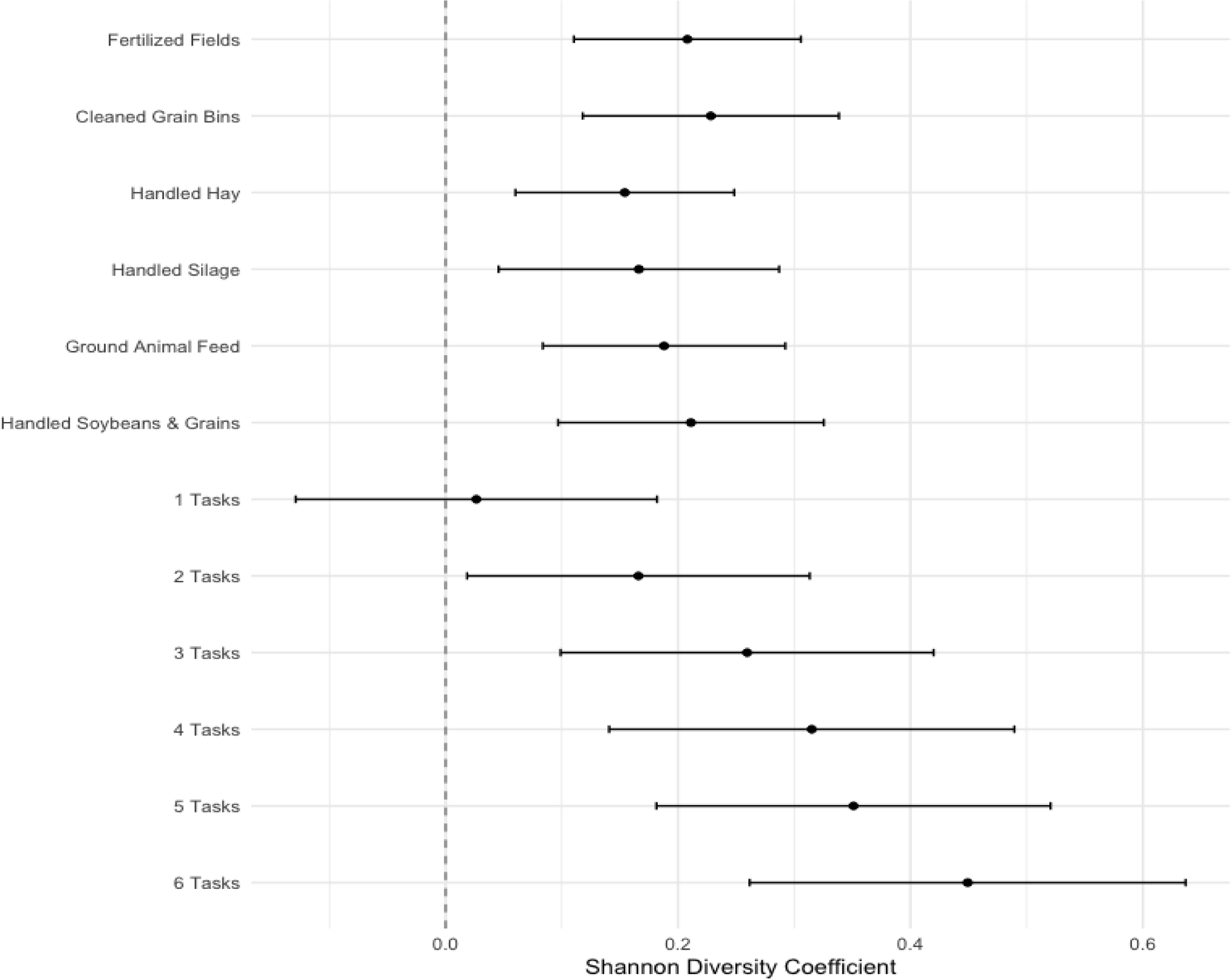
Work Tasks and Alpha Diversity in Home Dust Microbiome Samples. Coefficients derived from generalized linear regression models for each work tasks exposure, with Shannon alpha diversity index as the outcome, adjusted for age, smoking status, state of residence, asthma status, indoor pets, home condition, and home visit season. Error bars represent the 95% confidence intervals of the regression coefficients.

For beta-diversity (weighted UniFrac, **Table 5**) we found statistically significant differences in overall microbial composition for all work tasks (all p-values<0.001), with a moderate percent variance explained (R^2^ range 1.2%-3%). The sum of the work tasks accounted for higher explained variance (3%), followed by fertilizing fields (2%). Results for unweighted UniFrac distance were similar to the weighted metric for beta diversity (**Supplemental Table ST2**).

**Table 5:** Work Tasks and Weighted UniFrac Beta Diversity in Home Dust Microbiome Samples.

| Work Tasks | R<sup>2</sup> statistic | F p-value |
| --- | --- | --- |
| Fertilized Fields | 0.02 | 0.001 |
| Cleaned Grain Bins | 0.019 | 0.001 |
| Handled Hay | 0.016 | 0.001 |
| Handled Silage | 0.012 | 0.001 |
| Ground Animal Feed | 0.018 | 0.001 |
| Handled Soybeans & Grains | 0.014 | 0.001 |
| Job Tasks Sum | 0.03 | 0.001 |

Results from permutational multivariate analysis of variance (PERMANOVA) models for each work task exposure, with weighted UniFrac distance metric as the outcome, adjusted for age, smoking status, state of residence, asthma status, indoor pets, home condition, and home visit season. Presented is the R<sup>2</sup> value to quantify the percentage of variance explained by each exposure and the p-value for the F-statistic for compositional heterogeneity by exposure groups.

In analysis of individual microbial taxa, we found 21 taxa across 10 unique genera to be significantly differentially abundant by any work tasks (Figure 2). *Haloterrigena* was the most frequent differentially genera, with all six work tasks having higher abundance compared to not reporting any tasks. While most taxa (18) had increased abundance in relation to work tasks, 3 taxa within 3 genera had decreased abundance – *Thalassiosira* and *Fimbriimonas* for ground animal feed and *Oscillatoria* for handled hay. For the sum of the job tasks, only participants reporting 5 or 6 job tasks had significantly increased taxa abundance (*Sphaerobacter* and *Haloterrigena)* compared to participants’ reporting zero tasks (no differential taxa for 4 or fewer tasks).

**Figure 2:**
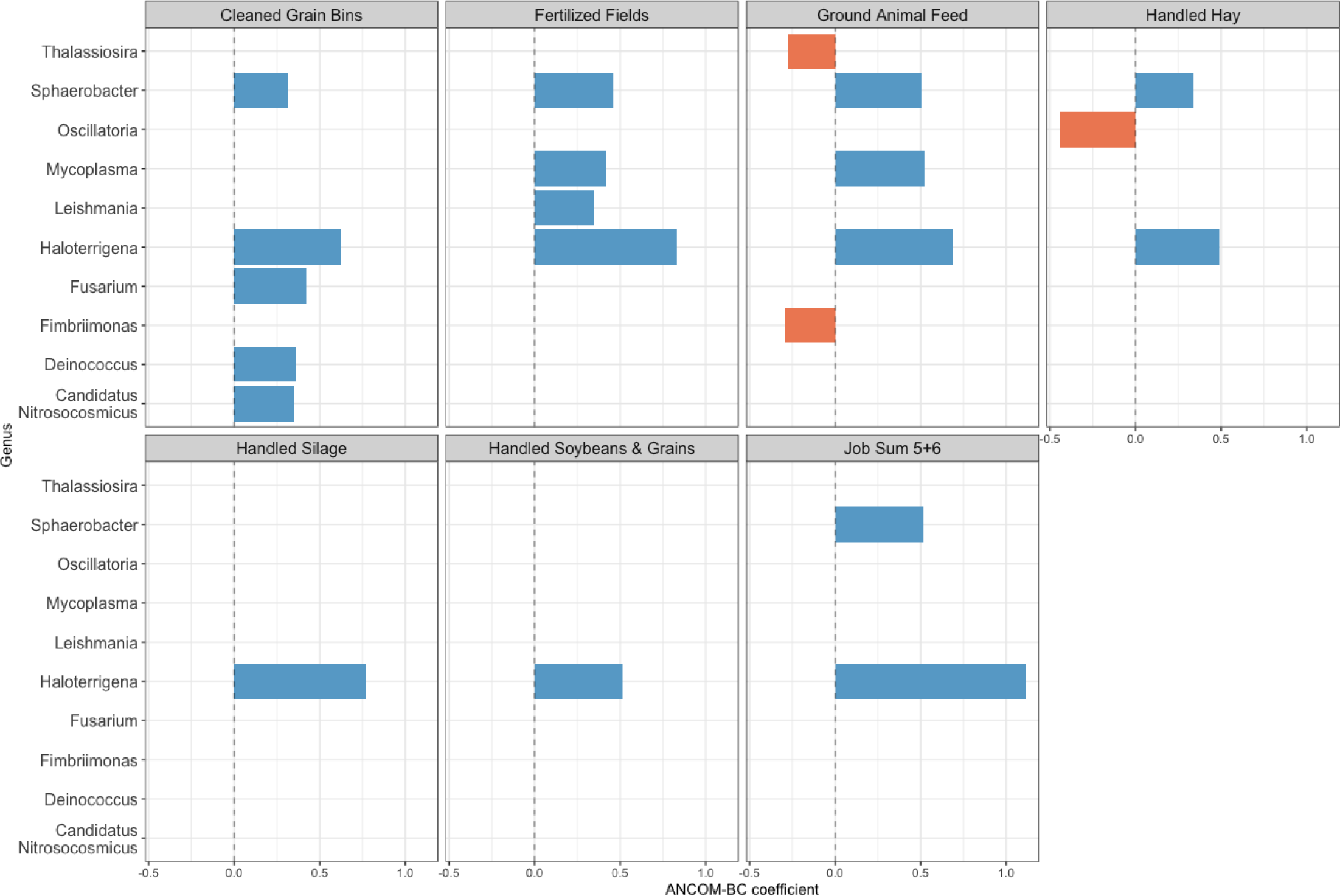
Work Tasks and Differentially Abundant Taxa in Home Dust Microbiome Samples. Figure shows the log fold-difference of the mean abundance of differentially abundant taxa (FDR p-value <0.05) by work tasks exposure groups, using the analysis of composition of microbiomes with bias correction (ANCOM-BC, v1.0.5) models.

### Varied Associations between Pesticides and the Home Dust Microbiome

Pesticide use was associated with home dust microbial diversity and composition, but to a lesser degree than work tasks. Of the 26 pesticide active ingredients, 19 had positive coefficients and 5 had negative coefficients for Shannon alpha diversity levels. Only current use of glyphosate and past use of lindane were statistically significant with confidence intervals above the null (glyphosate coefficient 0.18, 95% CI 0.02-0.36; lindane coefficient 0.11, 95% CI 0.03-0.21) (Figure 3). For beta compositional diversity (**Table 6**), 10 of the 19 currently used pesticides [acetochlor, atrazine, carbaryl, cyfluthrin, dicamba, glyphosate, permethrin, picloram, pyrethroid, 2,4-D] and all 7 banned pesticides were associated with altered weighted UniFrac beta diversity (range p-value 0.001-0.026). The explained variance for each pesticide (R^2^ range 0.4%-1.0%) was lower than that for any work task (R^2^ range 1.2%-3%). No pesticides were associated with unweighted UniFrac beta diversity (**Supplemental Table ST3**).

**Figure 3:**
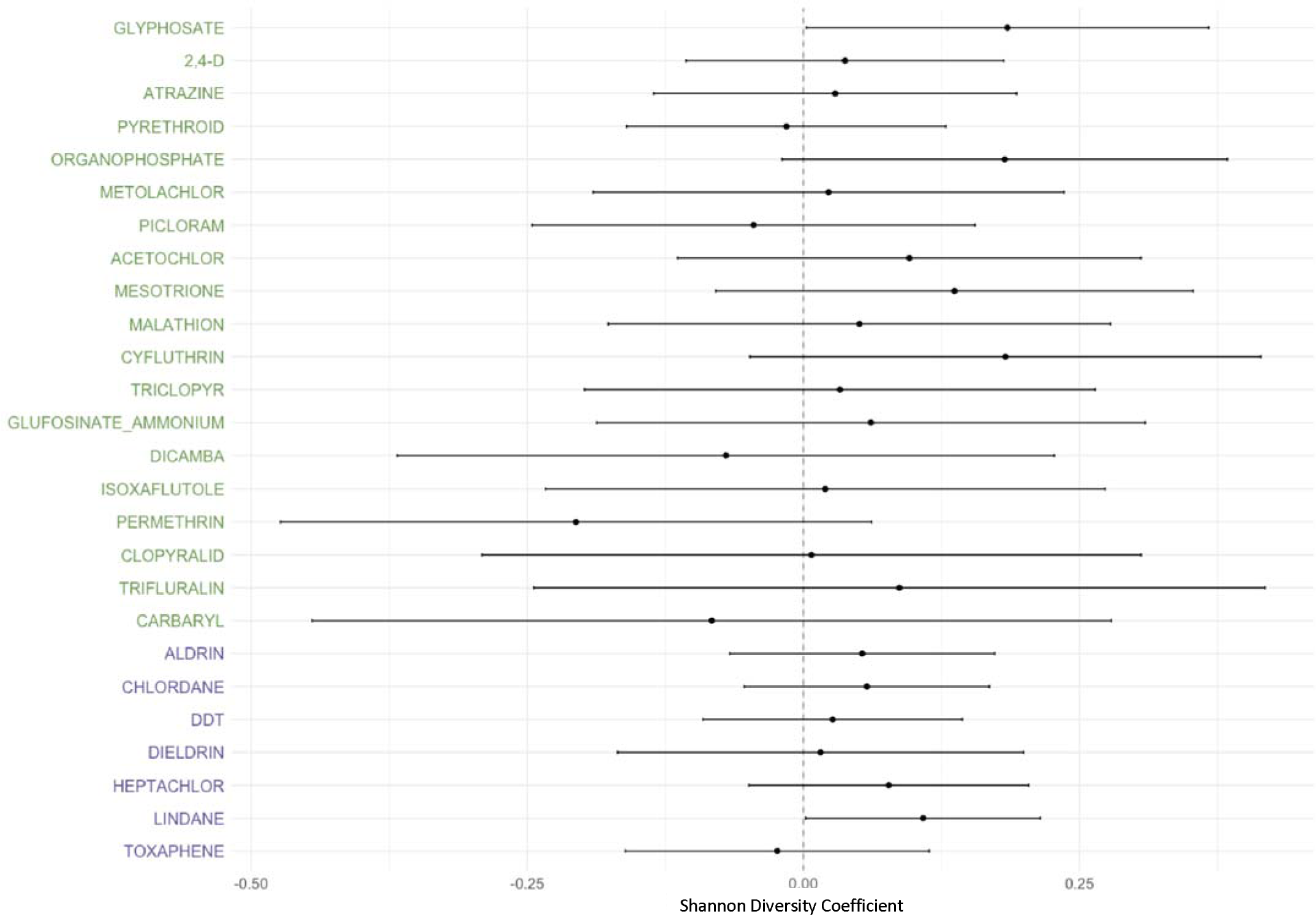
Pesticide Use and Alpha Diversity in Home Dust Microbiome Samples. Coefficients derived from generalized linear regression models for each pesticide, with Shannon alpha diversity index as the outcome, adjusted for age, smoking status, state of residence, asthma status, indoor pets, home condition, and home visit season. Green font indicates currently used pesticide and purple font indicates past use of banned pesticides. Error bars represent the 95% confidence intervals of the regression coefficients.

**Table 6:** Pesticide Use and Weighted UniFrac Beta Diversity in Home Dust Microbiome Samples.

| Current Pesticide | R2 statistic | F p-value | Banned Pesticide | R2 statistic | F p-value |
| --- | --- | --- | --- | --- | --- |
| ACETOCHLOR | 0.007 | 0.004 | ALDRIN | 0.007 | 0.012 |
| ATRAZINE | 0.009 | 0.001 | CHLORDANE | 0.007 | 0.01 |
| CARBARYL | 0.007 | 0.011 | DDT | 0.008 | 0.003 |
| CLOPYRALID | 0.004 | 0.586 | DIELDRIN | 0.006 | 0.019 |
| CYFLUTHRIN | 0.007 | 0.017 | HEPTACHLOR | 0.006 | 0.023 |
| DICAMBA | 0.01 | 0.001 | LINDANE | 0.007 | 0.014 |
| GLUFOSINATE_AMMONIUM | 0.004 | 0.341 | TOXAPHENE | 0.007 | 0.011 |
| GLYPHOSATE | 0.01 | 0.001 |  |  |  |
| ISOXAFLUTOLE | 0.005 | 0.216 |  |  |  |
| MALATHION | 0.005 | 0.131 |  |  |  |
| MESOTRIONE | 0.005 | 0.26 |  |  |  |
| METOLACHLOR | 0.004 | 0.458 |  |  |  |
| ORGANOPHOSPHATE | 0.005 | 0.186 |  |  |  |
| PERMETHRIN | 0.007 | 0.005 |  |  |  |
| PICLORAM | 0.006 | 0.026 |  |  |  |
| PYRETHROID | 0.01 | 0.001 |  |  |  |
| TRICLOPYR | 0.004 | 0.553 |  |  |  |
| TRIFLURALIN | 0.004 | 0.678 |  |  |  |
| 2,4-D | 0.007 | 0.003 |  |  |  |

Results from permutational multivariate analysis of variance (PERMANOVA) models for each pesticide exposure, with weighted UniFrac distance metric as the outcome, adjusted for age, smoking status, state of residence, asthma status, indoor pets, home condition, and home visit season. Presented is the p-value for the F-statistic for compositional heterogeneity by exposure groups and the $R^2$ value to quantify the percentage of variance explained.

In analyses of individual microbial taxa, there were 14 unique taxa belonging to 11 genera within 11 unique phyla that were differentially abundant in relation to at least one pesticide active ingredient (Figure 4). Current users of atrazine had decreased abundance of *Oscillatoria*, and current users of pyrethroid had decreased abundance of *Phocaeicola*, *Malassezia*, *Candidatus*, *Akkemansia*, and an unlabeled genus in the *Podoviridae* family. Increased abundances were seen for current 2,4-D and *Candidatus*, and past use of dieldrin, heptachlor, and lindane with *Treponema*, *Toxoplasma*, *Nitrospira*, *Haloterrigena*, *Phocaeicola*, *Malassezia*, and *Fusarium*. Overall, the ANCOM-BC coefficients representing log-fold changes in abundance for significant taxa were relatively small (range -0.75 to 0.67) compared to other microbiome studies (37).

**Figure 4:**
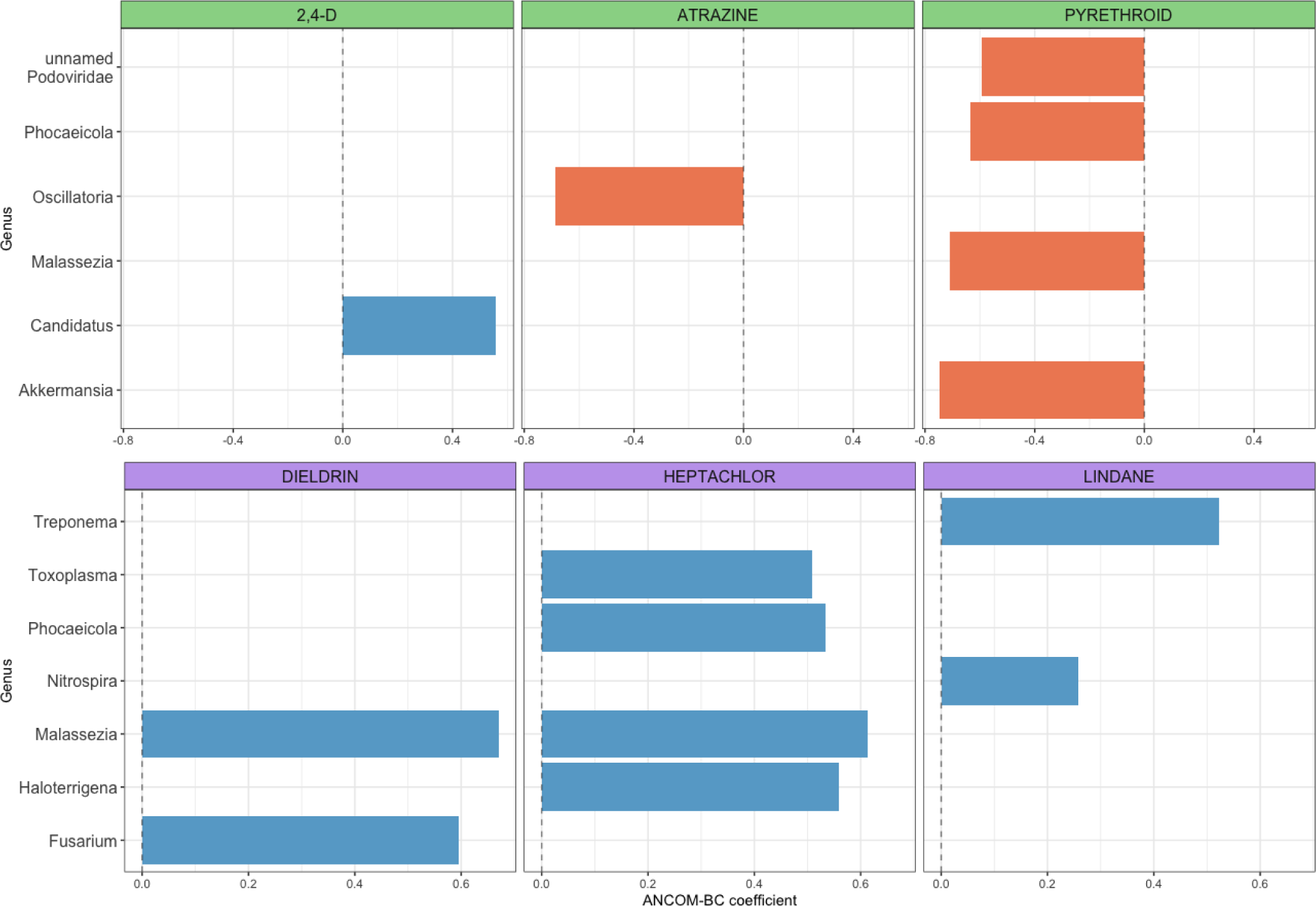
Pesticide Use and Differentially Abundant Taxa in Home Dust Microbiome Samples. Figure shows the log fold-difference of the mean abundance of differentially abundant taxa (FDR p-value <0.05) by pesticide exposure groups, using the analysis of composition of microbiomes with bias correction (ANCOM-BC, v1.0.5) models. Green facets are for currently used pesticide and purple facets are for past use of banned pesticides.

Associations of quantitative lifetime days of pesticide use with home dust microbiome are presented in **Supplemental Tables ST4-ST6**. For alpha diversity (**ST4**), carbaryl use days above the median was associated with increased within-sample diversity compared to never users (coefficient 0.13, 95% CI 0.003-0.25). Below the median days of use for dicamba (coefficient 0.13, 95% CI 0.003-0.26) and glyphosate (coefficient 0.22, 95% CI 0.07-0.37) were associated with increased within-sample diversity compared to never users. For both pesticides, days above the median had similar trends, but were not significant. We did not observe associations between alpha diversity and lifetime days of use for any of the banned pesticides. For beta diversity using weighted UniFrac beta diversity metric (**ST5**), lifetime days of use for 6 of the 19 current pesticides [acetochlor, atrazine, carbaryl, dicamba, glyphosate, metolachlor] and two of the banned pesticides [DDT, toxaphene] were associated with altered microbial composition. No lifetime days of pesticides use were associated with unweighted UniFrac metric. Twenty-nine unique taxa, within 16 unique genera and 16 unique phyla, were significantly differentially abundant across 9 pesticides by lifetime days of use (**ST6**).

### Independent Effect of Work Tasks and Pesticides on Home Microbial Diversity

To determine whether associations between work tasks and home dust microbiome alpha diversity were independent of pesticide use, we ran additional models adjusted for 1) the use of any pesticide in the past year and 2) for the two pesticide ingredients significantly associated with alpha diversity (glyphosate and lindane) (**Supplemental Table ST7**). Compared to models adjusting for demographics and home factors, adjustment for the use of any pesticide in past year had little effect on associations with the work tasks. Adjustment for use of lindane, a banned pesticide, also resulted in little effect size changes. However, because current use of glyphosate was highly prevalent at 86.2% and thus fairly strongly correlated with current work tasks, adjustment for glyphosate had a larger impact on the association. Associations were attenuated, with only 3 remaining statistically significant – fertilizing fields (coefficient 0.168, 95% CI 0.03-0.307), handling soybeans and grains (coefficient 0.281, 95% CI 0.048-0.515), and work tasks total (ordinal, coefficient 0.057, 95% CI 0.015-0.099). In analyses of categorical total number of work tasks, after adjusting for current glyphosate use, 5 or 6 total tasks remained significantly associated with alpha diversity (coefficients 0.349 and 0.384, 95% CI 0.006-0.691 and 0.023-0.746, respectively), which also had low correlation to glyphosate (tetrachoric correlation for 5 and 6 work tasks 0.16 and 0.14, respectively).

To determine whether associations between pesticide use and home dust microbiome alpha diversity were independent of work tasks, we ran additional models adjusted for work task total and specifically for fertilizing fields. Adjustment covariates were informed by independent associations between work tasks and alpha diversity after adjusting for pesticide use, and were less strongly correlated to pesticides (-0.23–0.6 and -0.28–0.61, respectively) (**Supplemental Table ST8**). Handling soybeans and grains was more strongly correlated with pesticide use (-0.31–0.82), hence we did not control for this task. No pesticide active ingredients were significantly associated with alpha diversity in models adjusting for either work tasks total or for fertilizing fields.

Given the complexity of the beta PERMANOVA and differential taxa abundance ANCOM-BC models, we did not conduct further analyzes to determine the independent effects of work tasks and pesticide use for these microbiome metrics.

## Discussion

This study is the first to assess the associations between agricultural work exposures and the worker’s home dust microbiome using metagenomic shotgun sequencing. We aimed to characterize farm occupational factors that influence the indoor built environment microbiome, a critical component to occupational and community health. We found that different work tasks were associated with altered diversity and composition of the microbes in participants’ homes. Further, we observed a dose-response relationship between microbial diversity and the number of farm work tasks performed. The use of certain pesticides, both current use and past use of banned pesticides, were associated with some differences in home dust microbiome, although associations were more modest than for the farm tasks. Overall, our findings suggest that the work that farmers perform in these facilities can impact the built environment of their homes, which could have implications for their own health and the health of any cohabitants in the home.

This research benefitted from the incorporation of in-depth details of participants’ reported farm work tasks. Previous studies evaluating potential hazards associated with farm work, particularly those integrating microbiome data, generally only assessed current job status and type of farm or commodity produced.(1, 38) However, our research showed robust associations between indoor microbiome and the tasks an individual performed on the farms, with a dose-response increase in microbial diversity linked to total number of tasks performed. All work tasks, including the sum of tasks preformed, were associated with altered beta composition metrics, with relatively high R^2^ explained variance compared to other studies.(19) All tasks were also associated with increased abundance of at least one microbe, the most frequent of which was *Haloterrigena*, a gram-negative soil-based bacteria.(39) Previous research has shown exposure to soil-based microbes in adult mouse models were associated with changes to the host microbiome, improved immune tolerance, and minimized allergic inflammation.(40)

Few studies have evaluated specific work tasks in relation to health outcomes or biomarkers of health. Within the Agricultural Health Study, specific farm work tasks were associated with systemic lupus (41), rheumatoid arthritis (42, 43), COPD (44), “famers’ lung” (specifically silage exposure) (45), wheeze (hay exposure) (46), stroke mortality (inversely associated with hay, grains, and silage) (47), non-Hodgkin lymphoid cancer (soybeans, grains, and hay) (48), and various injuries (49). Findings from other farming cohorts have found differences in lung cancer risk based on crop seedling and harvesting levels.(1) Additional studies have shown differences in adverse contaminant levels based on farm tasks, including mold (50) and black carbon.(51, 52) Research conducted in non-farming occupations have found differences in the microbiome of the host based on individual occupational exposures in salon workers (23), space station workers (53), firefighters (54), and janitorial staff.(55) However, to date, no studies have assessed farm work tasks associated with the microbiome of the workers at any host body site, or with the environmental microbiome. The findings from this work supports the need for careful evaluation of specific, detailed agricultural exposures to future occupational health studies.

In addition to specific work tasks, we explored the influence of participants’ use of pesticides on their home microbiome. The associations between pesticide use and dust microbiota were more varied than that for work tasks, in that not all pesticides were associated with changes to the home microbiome and some pesticides had opposite impacts on diversity levels and abundance of microbes, which might be expected due to the different mechanisms of action. We observed overall smaller R^2^ explained variance for beta diversity than work tasks, indicating pesticide use did not account for as much heterogeneity seen in the sample’s microbial composition as compared to work tasks, as well as lower log-fold changes for abundance of specific microbes compared to work tasks and previous literature using ANCOM-BC.(19) Pesticides that arose as related to the dust microbiome were current use of glyphosate, atrazine, 2,4-D, and pyrethroid, as well as past use of lindane. Atrazine and glyphosate, known bacterial degraders, have previously been associated with decreased diversity and decreased abundance of specific microbes in the soil microbiome.(56, 57)

Studies evaluating the effect of pesticides on the human microbiome have shown similarly heterogenous results to those in our study. Vindenes et al. found minimal associations between urinary pesticide metabolite concentration and oral microbiome metrics in a population-based study in Norway.(58) Conversely, Stanaway et al. found that the organophosphate insecticide azinphos-methyl was associated with changes in the oral microbiome in farmers in Yakima Valley, Washington USA, specifically decreased microbial diversity and reduced abundance of the *Streptococcus* genus.(21) While azinphos-methyl was not one of the active ingredients evaluated in our project, we did not see associations with reported use of organophosphates. Importantly, these research studies were evaluating urinary or serum concentrations of pesticide active ingredients, which generally reflect short-term exposure, whereas our study relied on self-report of pesticide use, an indicator of longer term exposure. Also, both prior studies focused on the oral microbiome and used 16S rRNA sequencing to characterize the microbiome, while we assessed the indoor environmental microbiota characterized by shotgun whole-genome sequencing, which can result in different exposure-outcome associations.(19) As ours is the first work to examine environmental microbiota in the homes of workers using advanced sequencing methodologies, it importantly adds to the growing literature of the effect of occupational chemical exposure on environmental microbiota, with inferences to health.

Our novel research benefits from a large sample population with detailed survey data on occupational exposures and characterization of the environmental microbiota using advanced techniques. However, this work does have limitations. First, work tasks were based on participant’s self-report, evaluated dichotomously (yes/no), as opposed to direct observation by a research technician. Yet, previous work in this population has shown these to correlate well with direct on-farm observations.(59) Similarly, pesticide use was assessed only by self-report. Nevertheless, biologic measurements of nonpersistent pesticides reflect only very recent use. Additionally, previous work in the target population has found strong accuracy of self-reported use of specific pesticides.(28, 60) Our analysis did not account for the rapidly growing area of pesticide and chemical mixtures, however current statistical mixtures methods do not accommodate binary exposure data.(61) Future development of mixtures methods and convenient code suitable for metagenomic analysis would be of interest. Although misclassification of farm exposure in this study is possible, we would expect this to be non-differential with respect to metagenomics and thus generally be a source of bias toward the null. In addition, our work task variables were highly correlated, making it challenging to assess their independent effect on the home dust microbiome. However, we were able to determine the magnitude of farming work by evaluating the total number of reported tasks, which can serve as a proxy for intensity of farming. A final limitation is that our dust sample was collected only from one location (participant bedroom) at one time and may not reflect the spatial and temporal heterogeneity in the environmental microbes found inside homes.(62) Again, this should be expected to reduce our ability to detect association and be a source of bias toward the null.

This work demonstrates that occupational exposures impact the microbiome inside farm workers’ homes as shown by altered diversity levels and abundance of specific microbes. Farm work tasks had more profound effects on home dust microbiota than use of pesticides. This is the first study to evaluate individual occupational exposures within farm work that associate with the indoor built environment microbiome. Our findings shed light on potential mechanistic pathways whereby occupational exposures can influence health through the role of the indoor microbiome and offers possible future intervention targets.

## Data Availability

Microbiome sequencing data are available at the Sequence Read Archive (SRA) under project number PRJNA975673 (https://www.ncbi.nlm.nih.gov/bioproject/PRJNA975673/). For metadata access, a data application will need to be approved by the Agricultural Health Study Executive Committee (www.aghealthstars.com).

## Supplemental Materials

See Supplemental Materials Table of Contents document for a list of the tables and figure referenced.

## Acknowledgments

We thank Dr. G. Ackermann for assistance with metadata curation, Dr. G. Humphrey for laboratory processing, Drs. R. Knight, A. González, and Q. Zhu for expert consultation, and the Center for Microbiome Innovation at the University of California San Diego for generating sequencing data. We thank Drs. F. Day of NIEHS for expert computational assistance and J. Hoppin (North Carolina State University, Raleigh, NC) for her important contribution to the Agricultural Lung Health Study during her tenure at NIEHS. We appreciate all the study participants for their contribution to this research.

## Data Sharing

**Occupational Farm Work Activities Influence Workers’ Indoor Home Microbiome** Dalton et. al.

## Notes

**Funding:** This work was supported by the Intramural Research Program of the National Institutes of Health (NIH), the National Institute of Environmental Health Sciences (NIEHS) (Z01-ES049030 and Z01-ES102385), the National Cancer Institute (Z01-CP010119B), and by American Recovery and Reinvestment Act funds.

### Competing Interest Statement

The authors have declared no competing interest.

### Funding Statement

This work was supported by the Intramural Research Program of the National Institutes of Health (NIH), the National Institute of Environmental Health Sciences (NIEHS) (Z01-ES049030 and Z01-ES102385), the National Cancer Institute (Z01-CP010119B), and by American Recovery and Reinvestment Act funds.

### Author Declarations

The Institutional Review Board at the National Institute of Environmental Health Sciences approved the study. Written informed consent was obtained from all participants.

